# Remdesivir for the treatment of patients hospitalized with COVID-19 receiving supplemental oxygen: a targeted literature review and meta-analysis

**DOI:** 10.1101/2022.03.04.22271706

**Authors:** Rachel Beckerman, Andrea Gori, Sushanth Jeyakumar, Jakob J. Malin, Roger Paredes, Pedro Póvoa, Nate Smith, Armando Teixeira-Pinto

**Author notes:** **Corresponding author:** Rachel Beckerman.

## Abstract

This network meta-analysis (NMA) assessed the efficacy of remdesivir in hospitalized patients with COVID-19 requiring supplemental oxygen. Randomized controlled trials of hospitalized patients with COVID-19, where patients were receiving supplemental oxygen at baseline and at least one arm received treatment with remdesivir, were identified. Outcomes included mortality, recovery, and no longer requiring supplemental oxygen. NMAs were performed for low-flow oxygen (LFO_2_); high-flow oxygen (HFO_2_), including NIV; or oxygen at any flow (AnyO_2_) at early (day 14/15) and late (day 28/29) time points. Six studies were included (N=5,245 patients) in the NMA. Remdesivir lowered early and late mortality among AnyO_2_ patients (risk ratio (RR) 0.52, 95% credible interval (CrI) 0.34-0.79; RR 0.81, 95%CrI 0.69-0.95) and LFO_2_ patients (RR 0.21, 95%CI 0.09-0.46; RR 0.24, 95%CI 0.11-0.48); no improvement was observed among HFO_2_ patients. Improved early and late recovery was observed among LFO_2_ patients (RR 1.22, 95%CrI 1.09-1.38; RR 1.17, 95%CrI 1.09-1.28). Remdesivir also lowered the requirement for oxygen support among all patient subgroups. Among hospitalized patients with COVID-19 requiring supplemental oxygen at baseline, use of remdesivir compared to best supportive care is likely to improve the risk of mortality, recovery and need for oxygen support in AnyO_2_ and LFO_2_ patients.

## Introduction

Infection with SARS-CoV-2 can cause coronavirus disease 2019 (COVID-19) and, in severe cases, patients may present with acute respiratory distress syndrome or septic shock with multiple organ failure.^1^ Compared to seasonal influenza, patients with COVID-19 are more likely to be hospitalized, need intensive care, have a longer duration of hospitalization, and die in hospital.^2^ Further, severe COVID-19 patients are at a higher risk for hospital-acquired infections, namely ventilator-associated pneumonia, and have increased rates of multiorgan dysfunction.^3-5^

Remdesivir (GS-5734) is a ribonucleic acid (RNA)-dependent RNA polymerase inhibitor that was identified early as a promising therapeutic candidate for COVID-19 due to its broad inhibitory activity against RNA viruses such as the Middle East Respiratory Syndrome,^6^ and acts as a nucleoside analog, inhibiting the RNA-dependent RNA polymerase of SARS-CoV-2.^7^ Clinical trials were initiated in 2020 to evaluate the safety and efficacy of remdesivir, among other drugs, as treatments for COVID-19. These included the National Institute of Allergy and Infectious Diseases Adaptive COVID-19 Treatment Trials (ACTT-1 and ACTT-2) which assessed the impact of remdesivir, alone or in combination, on time to recovery; ^8,9^ and the World Health Organization (WHO)-led SOLIDARITY trial which compared remdesivir, lopinavir/ritonavir, lopinavir/ritonavir with interferon-B1a and chloroquine or hydroxychloroquine on mortality.^10^ ACTT-1, the pivotal double-blind, randomized, placebo-controlled trial, found that treatment with remdesivir resulted in shorter median recovery time compared to those who received placebo; post-hoc-analyses among low-flow oxygen patients suggested remdesivir resulted in a 70% reduction in mortality.^8^ While results in SOLIDARITY were not stratified by supplemental oxygen needs, there was a trend towards a clinical benefit of remdesivir for patients on oxygen versus patients who were ventilated.^10^ Despite this, following the interim results of SOLIDARITY^10^, the WHO concluded that remdesivir had little or no effect on hospitalized patients with COVID-19, as determined by overall mortality.

Given the ongoing global emergency of the disease and rapid viral evolution of SARS-CoV-2, effective and safe treatments for patients with COVID-19 are still urgently needed. Multiple meta-analyses have been conducted in order to determine the clinical significance of remdesivir for patients with COVID-19.^11-21^ However, the role of remdesivir by supplemental oxygen needs is not yet fully understood. This review and meta-analysis includes previously unavailable data to evaluate the efficacy of remdesivir in hospitalized COVID-19 patients requiring low- and/or high-flow oxygen on key endpoints of interest.

## Materials and Methods

### Study Design

This study followed the preferred reporting items for systematic reviews and meta-analysis (PRISMA) statement for study design (**Table S1 (Supplementary Materials)**.^22^

### Outcomes

Key outcomes of interest were mortality; recovery (defined as either recovery from COVID-19 or discharge from hospital, and was assumed to be interchangeable despite varying definitions of recovery across trials); no longer requiring supplemental oxygen; or progressing to NIV or invasive mechanical ventilation (IMV). Outcomes were stratified by the population for which remdesivir has been conditionally approved to treat COVID-19 by the European Medicines Agency (EMA): patients with pneumonia requiring supplemental oxygen (low- or high-flow oxygen or other NIV) at the start of treatment. These were defined as oxygen at any flow, high-flow oxygen (which included, in some trials, patients receiving non-invasive ventilation [NIV]), or low-flow oxygen. Patients in trials who were on NIV at baseline (included in this analysis when grouped in an ordinal group that included patients with high-flow oxygen or NIV) and remained on NIV, were considered to have progressed as they did not recover.

### Search Strategy and Inclusion Criteria

A targeted search was conducted over three months (February to April, 2021) to identify relevant materials in MEDLINE (PubMed), medRxiv, EMBASE and Cochrane Trials (**Table S2, Supplementary Materials**). Inclusion criteria for studies were randomized controlled trials (either published or in pre-print) that enrolled patients hospitalized requiring supplemental oxygen at baseline. Patients in at least one arm of the trial must have been treated with remdesivir and the trial had to report on at least one outcome of interest on day 14/15 or day 28/29. In trials that reported on both patients who did and did not receive supplemental oxygen, only those patients who required supplemental oxygen at baseline were included.

### Data Extraction & Risk of Bias Evaluation

Data extraction was done by one researcher. Outcomes reported at different time points were considered equivalent: day 14 to day 15 and 28 to day 29. One study reported outcomes at day 24^23^ and it was assumed to be equivalent to the day 28/29 time point. Risk of bias was evaluated using the revised risk of bias assessment for randomized controlled trials tool by one member of the research team.^24^

### Statistical analysis

Given the lack of statistical difference for 5-versus 10-day treatment of remdesivir in previous meta-analyses^13,25,26^, this analysis aggregated 5- and 10-day treatment. All outcomes were analyzed using standard Bayesian techniques, adapting previously validated methods.^27,28^ A Bayesian network meta-analysis, using a generalized linear model (with binomial likelihood and log link) for each outcome, was implemented using BUGSnet. Non-informative prior distributions were used for all parameters (**Table S3, Supplementary Materials**).^29^ The Markov chain Monte Carlo simulations were specified as a burn-in of 50,000 iterations followed by 100,000 iterations with 10,000 adaptations. Trace plots and density plots were used to evaluate convergence graphically. Both fixed and random effect models have been utilized in prior remdesivir meta-analyses^11-19^. While model fits were similar for fixed and random effects (**Table S4, Supplementary Materials**), given the small number of studies included in the analysis, a fixed effects model was selected as the base case. Results of the random effects model are included in the **Supplementary Materials**. Consistency within the network was assessed using the individual data points’ posterior mean deviance contributions for the consistency model versus the inconsistency model, following recommendations.^30^ Results are presented as risk ratios (RR) between treatment and best supportive care with forest plots. Surface under the curve cumulative ranking probabilities (SUCRA) plots are also presented to show the ranking of treatments. Credible intervals (CrI) of 95% were used for inference. All data analyses were performed using Microsoft Excel (2019) and the R statistical package. Scenario analyses were performed to evaluate the robustness of the models’ results. The first included data from the SIMPLE-Severe trial^25^, via a matched historical control study^31^, where remdesivir was compared to a control arm of a retrospective cohort of patients with severe COVID-19 (via inverse probability weighted multiple logistic regression). The second scenario analysis excluded ACTT-2 from the analysis, thereby only including comparisons of remdesivir versus standard of care. The third scenario analysis explored 5- and 10-day treatment with remdesivir, separately, versus best supportive care.

## Results

### Search and study selection process

A total of 2,634 unique studies were retrieved from the databases and 42 studies were retained for full-text review; a further 36 were excluded (**Figure 1**). While SIMPLE-Moderate^26^ did not report results stratified by the EMA population, the authors were contacted and were able to provide the appropriate data; thus, this study was included. Further, following construction of the networks (**Figure S1, Supplementary Materials**), it was determined that when aggregating the 5- and 10-day treatment arms, SIMPLE-Severe^25^ could no longer be connected to the network and was thus excluded from the base case analysis. Therefore, in the base case, a total of six studies were entered into the meta-analysis ^8-10,23,26,32^ (**Figure 1**).

**Figure 1.**
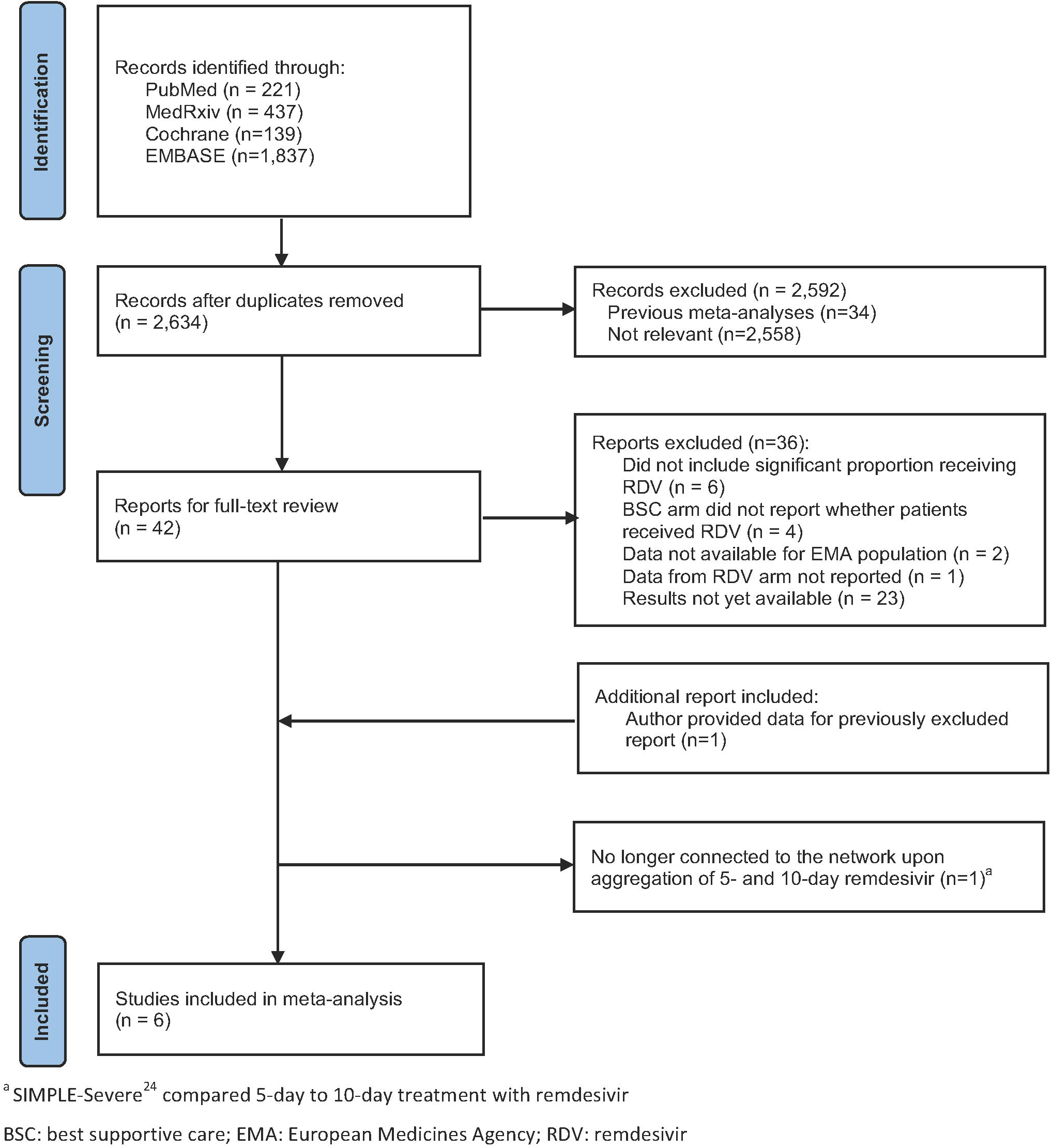
PRISMA study selection flow diagram.

### Risk of bias assessment results

Risk of bias, as assessed by the revised risk of bias assessment tool is presented in **Table S5 (Supplementary Materials)**

### Characteristics of studies included in the analysis

Characteristics of the included studies are presented in **Table 1**. Patient characteristics from the included studies are presented in **Table 2**. Treatment with remdesivir was consistently administered intravenously as 200 mg on day 1 followed by 100 mg for either 4 or 9 days. Across all trials, all patients could receive best supportive care in all treatment arms.

**Table 1.** Characteristics of randomized controlled trial studies included

| Study<br>Country Author | Design | Phase | Number<br>randomized | Follow-up<br>period | Primary endpoint | Arm 1 | Arm 2 | Arm 3 |
| --- | --- | --- | --- | --- | --- | --- | --- | --- |
| ACTT-1<br>Multi-country<br>Beigel<sup>8</sup> | Double-blind,<br>placebo-controlled | 3 | 1,062 | 29 days | Time to recovery | RDV by IV as 200 mg on day 1, followed by 100 mg on days 2 - 10 or until discharge or death plus supportive care | Matching placebo plus supportive care | N/A |
| ACTT-2<br>Multi-country<br>Kalil<sup>9</sup> | Double-blind,<br>placebo-controlled | 3 | 1,033 | 29 days | Time to recovery | Baricitinib as 4 mg daily for 14 days or until discharge plus RDV by IV as 200 mg on day 1, followed by 100 mg on days 2 -10 or until discharge or death plus supportive care | RDV by IV as 200 mg on day 1, followed by 100 mg on days 2 - 10 or until discharge or death plus placebo plus supportive care | N/A |
| Hubei<br>China<br>Wang<sup>32</sup> | Double-blind,<br>placebo-controlled | 3 | 236 | 28 days | Time to clinical improvement up to day 28 | RDV by IV as 200 mg on day 1, followed by 100 mg on days 2 -10 | Matching placebo | N/A |
| SOLIDARITY<br>Multi-country<br>WHO Solidarity Consortium<sup>10</sup> | Open-label | 3 | 5,475* | 28 days | In-hospital mortality | RDV by IV as 200 mg on day 1, followed by 100 mg on days 2 -10 plus supportive care | Standard of care according to local hospital | N/A |
| SIMPLE-Moderate<br>Multi-country<br>Spinner<sup>26</sup> | Open-label | 3 | 596 | 28 days | Clinical status on day 11 | RDV by IV as 200 mg on day 1, followed by 100 mg on days 2 -4 plus supportive care | RDV by IV as 200 mg on day 1, followed by 100 mg on days 2 -10 plus supportive care | Best supportive care |
| Mahajan<br>India<br>Mahajan<sup>23</sup> | Open-label | NR | 82 | 24 days | Improvement in clinical outcome | RDV by IV as 200 mg on day 1, followed by 100 mg on days 2 -5 plus supportive care | Standard of care | N/A |

\*Remdesivir and control arm only
IV: intravenous; N/A: not applicable; NIV: non-invasive ventilation; NR: not reported; RDV: remdesivir;

**Table 2.**
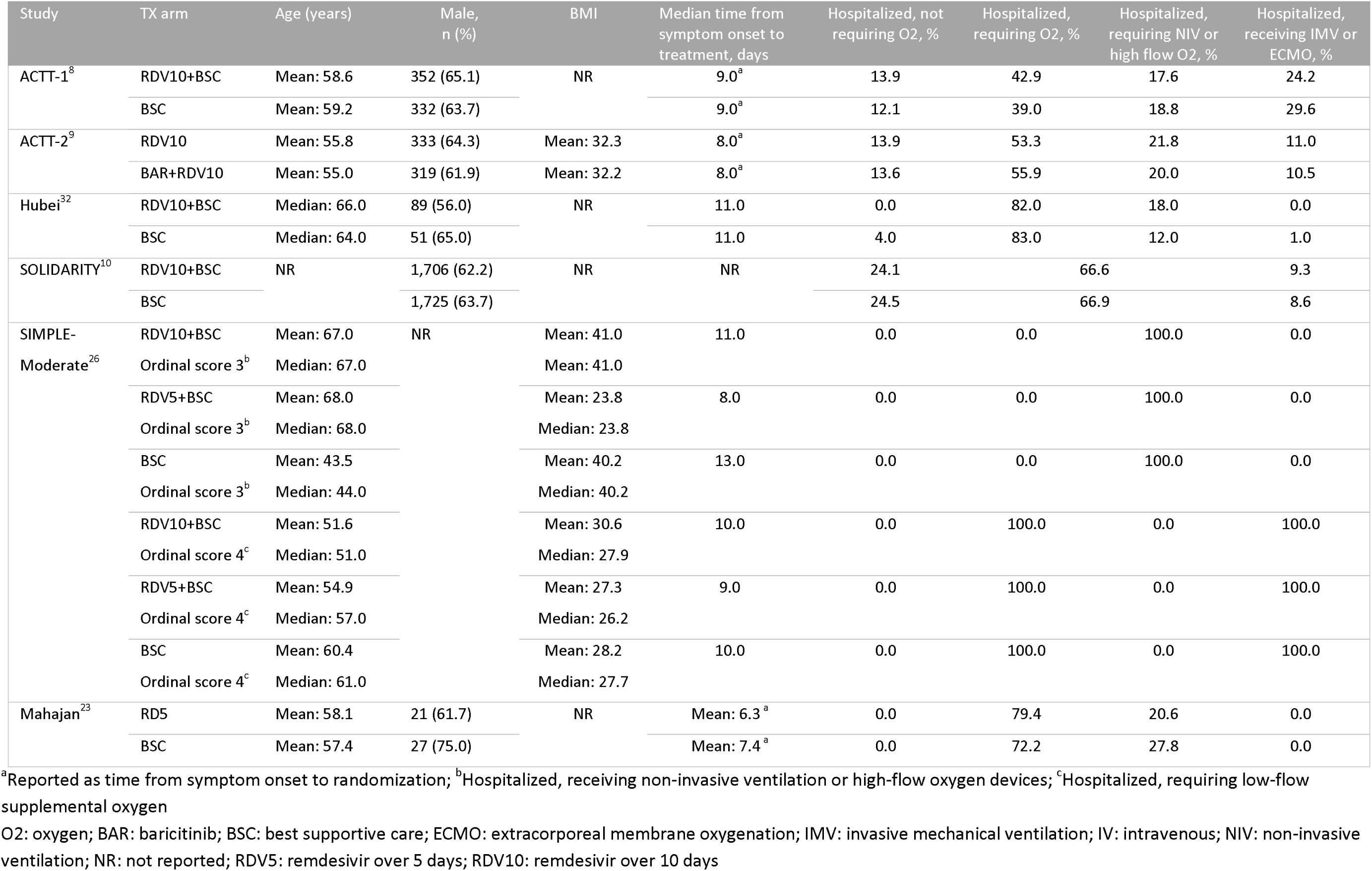
Patient characteristics of included studies included

### Outcomes

A summary of the outcomes included in the meta-analysis, stratified by subpopulation, are presented in **Table 3**. Earlier mortality included assessment at day 14^9,26,32^ or day 15^8^; later mortality included assessment at day 28 ^9,10,26,32^ or day 29^8^. Mahajan^23^ assessed outcomes at day 24 and was included with the later assessment. Five studies reported recovery or discharges at both the early (day 14/15) and later (day 28/29) time point ^8-10,26,32^; Mahajan^23^ assessed discharges at day 24 and was considered with the later assessment. There was insufficient data to analyze either no longer requiring oxygen support or progressing to NIV or IMV at the later time point of assessment; thus, only the early timing of assessment for these outcomes is reported.

**Table 3.** Summary of outcomes by oxygen flow requirements

| Study | Treatment arm | Mortality |  | Recovery or discharges |  | No longer requiring O2 | Requiring NIV<sup>a</sup> | Requiring IMV<sup>a</sup> |
| --- | --- | --- | --- | --- | --- | --- | --- | --- |
|  |  | Early | Later | Early | Later | Early | Early | Early |
| Any non-invasive oxygen flow |  |  |  |  |  |  |  |  |
| ACTT-1<sup>8</sup> | RDV10+BSC | 20/327 | 28/327 | 206/327 | 263/327 | 235/327 | 17/327 | 29/327 |
|  | BSC | 38/301 | 45/301 | 157/301 | 217/301 | 174/301 | 18/301 | 41/301 |
| ACTT-2<sup>9</sup> | RDV10 | 4/391 | 9/389 | 293/391 | 344/391 | 314/391 | 16/391 | 23/391 |
|  | BAR+RDV10 | 12/391 | 25/389 | 258/389 | 316/389 | 266/389 | 17/389 | 47/389 |
| Hubei<sup>32</sup> | RDV10+BSC | 15/153 | 22/158 | 39/153 | 92/150 | 60/153 | 13/153 | 4/153 |
|  | BSC | 7/78 | 10/78 | 18/78 | 45/77 | 28/78 | 8/78 | 7/78 |
| SOLIDARITY<sup>10</sup> | RDV10+BSC | NR | 192/1,828 | 1,234/1,828 | 1,507/1,828 | NR | NR | NR |
|  | BSC | NR | 219/1,811 | 1,241/1,811 | 1,468/1,811 | NR | NR | NR |
| Mahajan<sup>23</sup> | RD5 | NR | 5/34 | NR | 2/34 | NR | NR | NR |
|  | BSC | NR | 3/36 | NR | 3/34 | NR | NR | NR |
| SIMPLE-Moderate<sup>26</sup> | RDV10+BSC | 0/24 | 0/24 | 22/24 | 23/24 | 23/24 | 0/24 | 0/24 |
|  | RDV5+BSC | 0/31 | 0/31 | 24/31 | 28/31 | 27/31 | 2/31 | 0/31 |
|  | BSC | 4/38 | 4/38 | 23/38 | 29/38 | 26/38 | 2/38 | 3/38 |
| Low flow oxygen<sup>b</sup> |  |  |  |  |  |  |  |  |
| ACTT-1<sup>8</sup> | RDV10+BSC | 7/232 | 9/232 | 166/232 | 206/232 | 183/232 | 5/232 | 13/232 |
|  | BSC | 21/203 | 25/203 | 124/203 | 156/203 | 137/203 | 7/203 | 21/203 |
| ACTT-2<sup>9</sup> | RDV10 | 3/288 | 5/288 | 236/288 | 262/288 | 250/288 | 9/288 | 8/288 |
|  | BAR+RDV10 | 4/276 | 12/276 | 217/276 | 243/276 | 224/276 | 1/276 | 19/276 |
| SIMPLE-Moderate<sup>26</sup> | RDV10+BSC | 0/23 | 0/23 | 22/23 | 22/23 | 23/23 | 0/23 | 0/23 |
|  | RDV5+BSC | 0/29 | 0/29 | 24/29 | 28/29 | 27/29 | 0/29 | 0/29 |
|  | BSC | 4/36 | 4/36 | 22/36 | 27/36 | 25/36 | 1/36 | 3/36 |
| High flow oxygen<sup>c</sup> |  |  |  |  |  |  |  |  |
| ACTT-1<sup>8</sup> | RDV10+BSC | 13/95 | 19/95 | 40/95 | 57/95 | 52/95 | 12/95 | 16/95 |
|  | BSC | 17/98 | 20/98 | 33/98 | 61/98 | 37/98 | 11/98 | 20/98 |
| ACTT-2<sup>9</sup> | RDV10 | 1/103 | 5/113 | 57/103 | 82/103 | 64/103 | 7/103 | 15/103 |
|  | BAR+RDV10 | 7/103 | 13/113 | 41/113 | 73/113 | 44/113 | 16/113 | 28/113 |
| SIMPLE-Moderate<sup>26</sup> | RDV10+BSC | 0/1 | 0/1 | 0/1 | 1/1 | 0/1 | 0/1 | 0/1 |
|  | RDV5+BSC | 0/2 | 0/2 | 0/2 | 0/2 | 0/2 | 2/2 | 0/2 |
|  | BSC | 0/2 | 0/2 | 1/2 | 2/2 | 1/2 | 1/2 | 0/2 |

<sup>a</sup>Or worse; <sup>b</sup>Low-flow oxygen defined as either hospitalized and requiring any supplemental oxygen or hospitalized requiring low-flow supplemental oxygen, depending on the study; <sup>c</sup>High-flow oxygen defined as hospitalized and requiring non-invasive ventilation or use of high-flow oxygen devices, depending on the study
BAR: baricitinib; BSC: best supportive care; IMV: invasive mechanical ventilation; NIV: non-invasive ventilation; NR: not reported; O2: oxygen; RDV5: remdesivir over 5 days; RDV10: remdesivir over 10 days

Overall, there was a lack of evidence to suggest inconsistency within the networks (**Figures S2-S6, Supplementary Materials**).

### Mortality

Treatment with remdesivir was superior in lowering the risk of mortality among patients receiving any supplemental oxygen (early assessment RR [95% CrI]: 0.52 [0.34, 0.79]; late assessment RR: 0.81 [0.69, 0.95]) and those receiving only low-flow oxygen at both the early (RR: 0.32 [0.09, 0.46]) and late assessment (RR: 0.24 [0.11, 0.48]) (**Figure 2**). Treatment with remdesivir, however, did not lower the risk of mortality among patients receiving high-flow oxygen at either the early or later endpoint assessment (**Figure 2**). Results were similar for treatment with remdesivir in combination with baricitinib, with the exception of mortality at the early assessment among low-flow oxygen patients. Treatment with remdesivir (with or without baricitinib) was ranked superior to the standard of care across all patient subgroups at both the early and later assessment for the mortality endpoint (**Table S6, Supplementary Materials**).

**Figure 2.**
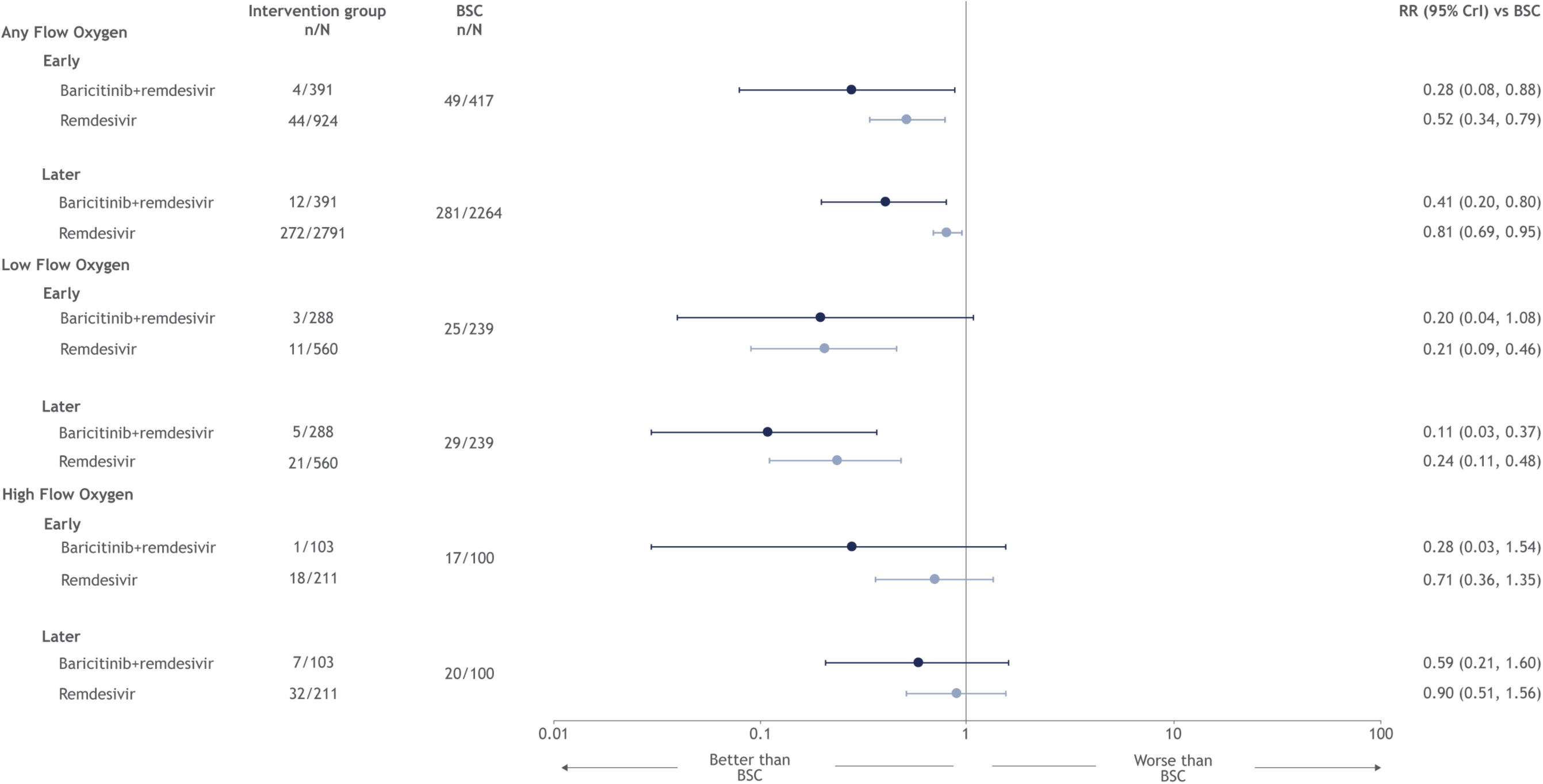
Forest plot for mortality endpoint, by type of non-invasive oxygen support.

### Recovery

Treatment with remdesivir was superior in improving recovery among those on low-flow oxygen at both the early (RR: 1.22 [1.09, 1.38]) and later (RR: 1.17 [1.09, 1.28]) assessment; treatment with remdesivir did not improve recovery in patients receiving any supplemental oxygen or on high-flow oxygen (**Figure 3**). Treatment with remdesivir in combination with baricitinib was superior in improving recovery in all patients, with the exception of those on high-flow oxygen at the later assessment. Treatment with remdesivir was ranked superior to standard of care across all patient subgroups at both the early and later assessment for the recovery endpoint (**Table S6, Supplementary Materials**).

**Figure 3.**
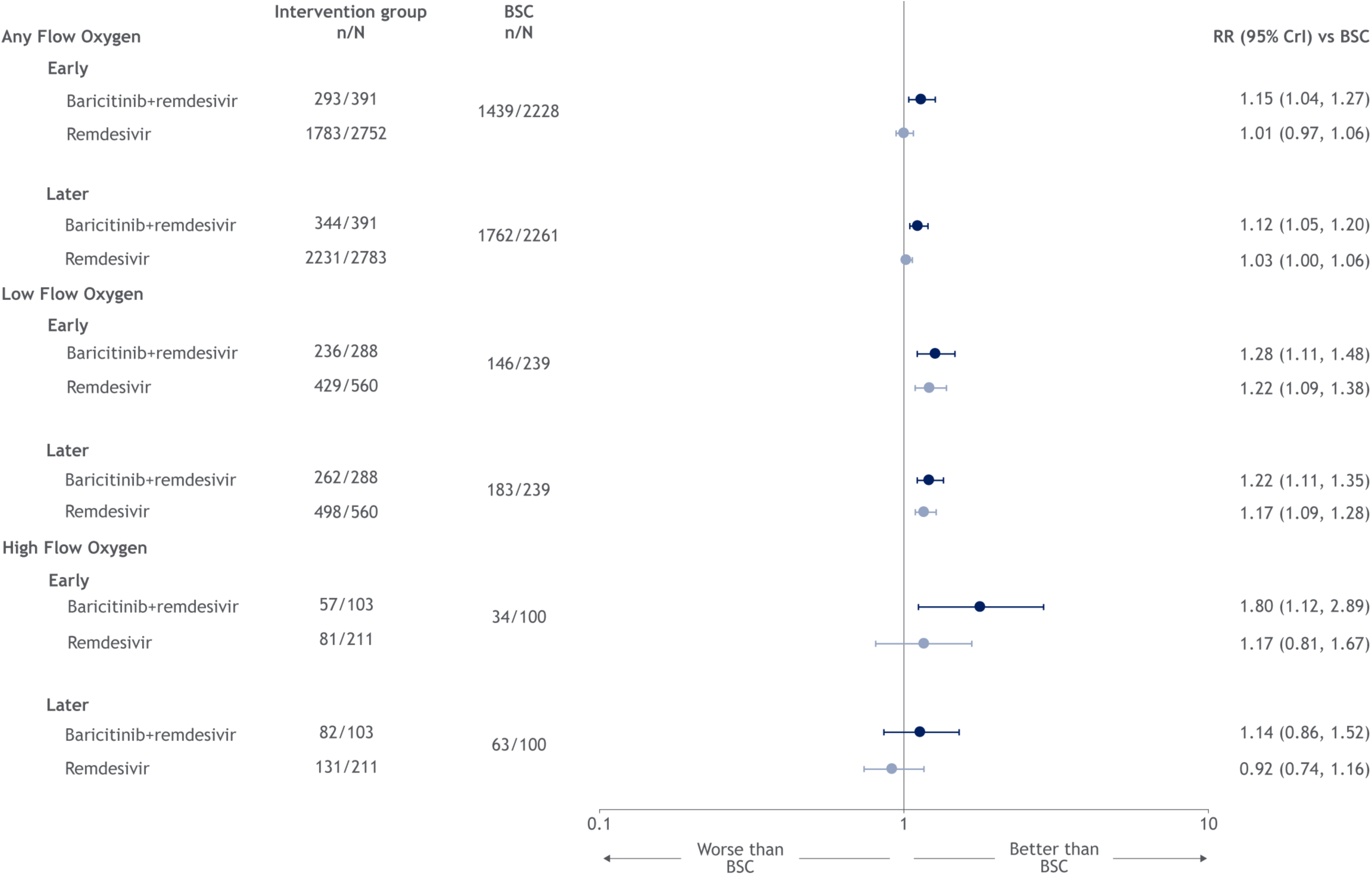
Forest plot for recovery endpoint, by type of non-invasive oxygen support.

### No longer requiring oxygen support

Treatment with remdesivir increased the likelihood of no longer requiring oxygen support among all patient subgroups at day 14 **(Figure 4)**. Among patient subgroups, RR (95% CrI) varied from 1.22 (1.11, 1.35) among low-flow oxygen patients to 1.37 (1.01, 1.88) among high-flow oxygen patients. Treatment with remdesivir was ranked superior to the standard of care for no longer requiring oxygen support endpoint across all patient subgroups for the oxygen support endpoint (**Table S6, Supplementary Materials**). Similar results were observed for remdesivir in combination with baricitinib (**Figure 4**).

**Figure 4.**
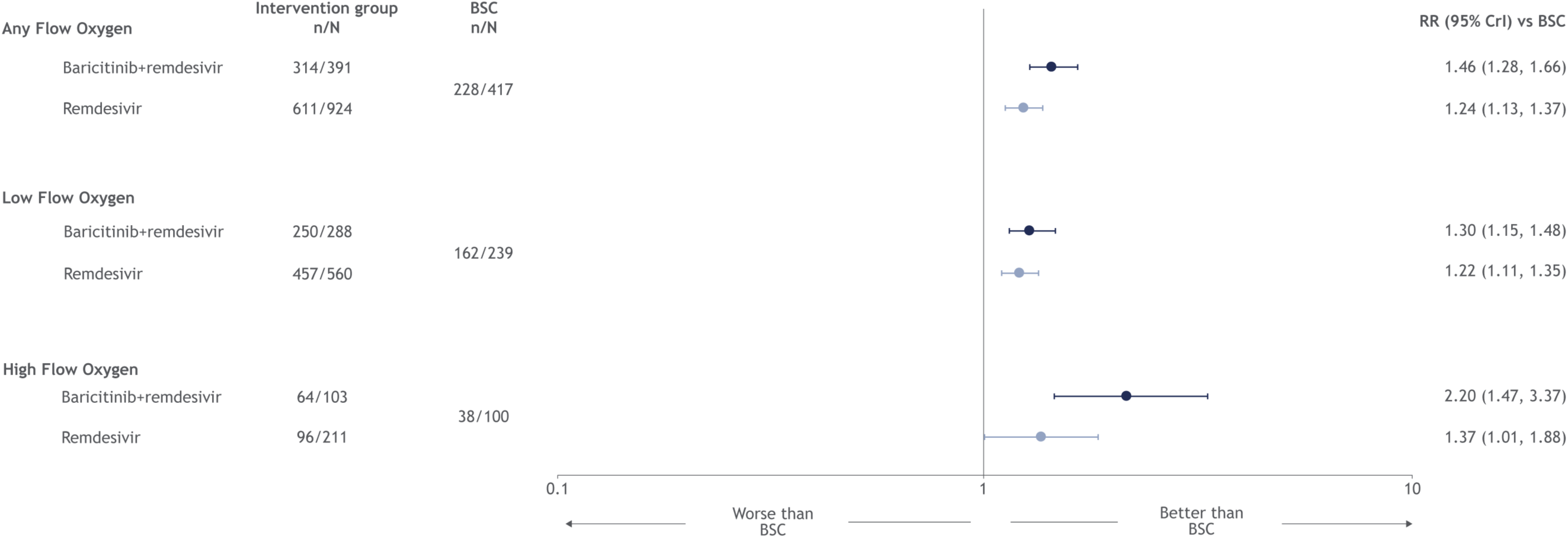
Forest plot for free from oxygen support endpoint, by type of non-invasive oxygen support.

### Progressing to NIV or IMV

Treatment with remdesivir lowered the risk of progression to NIV or worse among patients on any supplemental oxygen (RR: 0.56 [0.47, 0.67]) and low-flow oxygen (RR: 0.37 [0.23, 0.56]) and lowered the risk of progression to IMV or worse among patients on any supplemental oxygen (RR: 0.54 [0.41, 0.71]) and high-flow oxygen (RR: 0.34 [0.20, 0.54]) (**Figure 5)**. For both NIV and IMV, treatment with remdesivir was ranked superior to the standard of care across all patient subgroups (**Table S6, Supplementary Materials**). Treatment with remdesivir in combination with baricitinib lowered the risk of progression to NIV or worse, or IMV or worse, across all patient subgroups at both the early and late time assessment.

**Figure 5.**
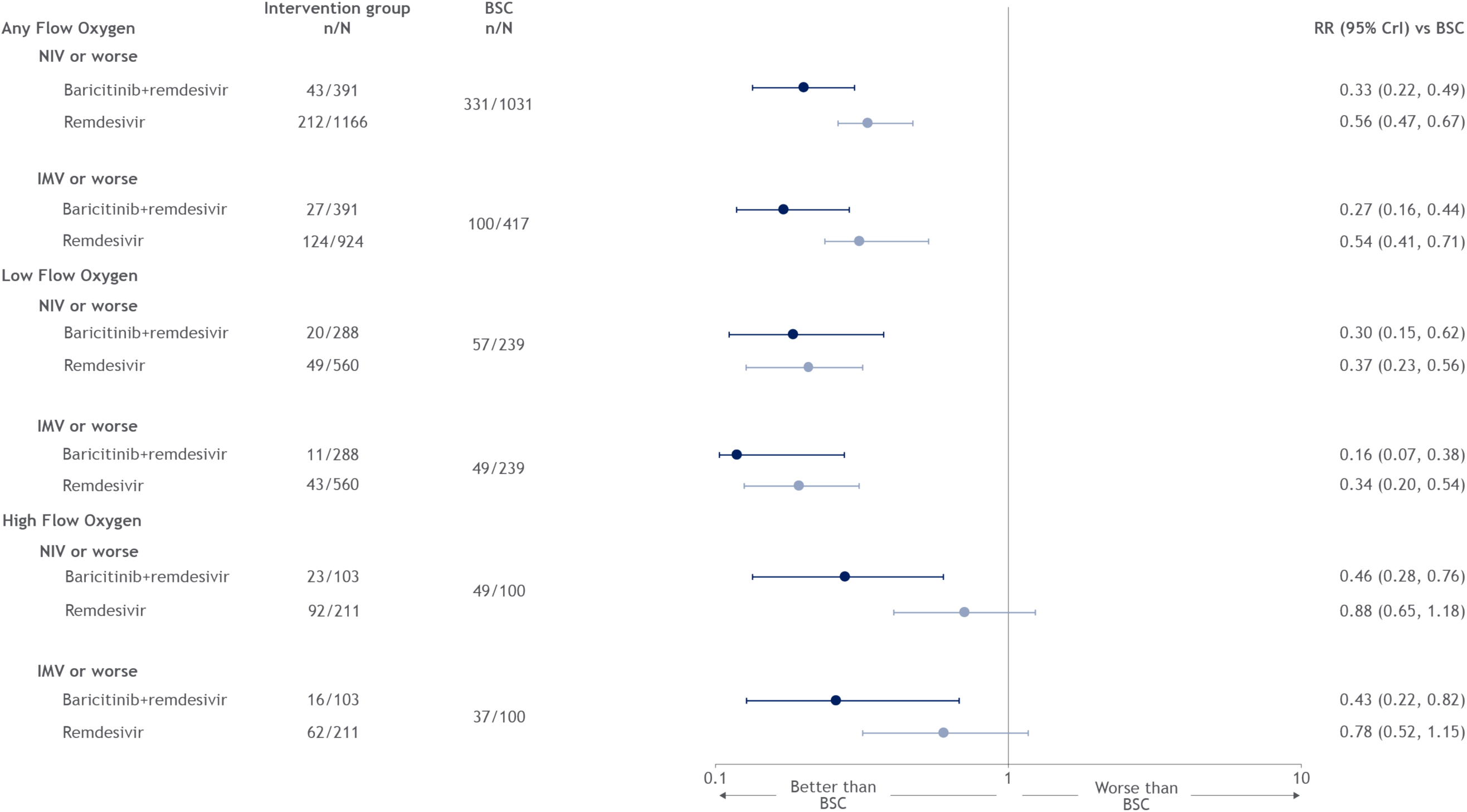
Forest plot for need for non-invasive ventilation or invasive medical ventilation support endpoint, by type of non-invasive oxygen support.

### Scenario analyses

When treatment with remdesivir was disaggregated for 5- and 10-days, results were similar to the base case analysis (**Figure S7**). However, given the few patients available to the network for 5-day remdesivir, effect estimates are uncertain as reflected by the wide credible intervals.

ACTT-2 compared treatment with remdesivir to remdesivir in combination with baricitinib. When ACTT-2 was excluded from the network, results for remdesivir were similar to the base case analysis. Remdesivir significantly decreased mortality among patients on any flow and on low-flow oxygen (**Figure S8**). Results for the endpoints recovery, no longer requiring oxygen support and progressing to more intensive oxygen support (either NIV or IMV or worse, depending on baseline oxygen status) were similar to the base case analysis (**Figures S9 – S11**). For all endpoints for the low- and high-flow oxygen subgroups, only ACTT-1 and SIMPLE-Moderate informed the analyses.

SIMPLE-Severe^25^ only reported outcomes for the early time assessment, therefore, this scenario analysis only explored outcomes at day 14/15. When data from SIMPLE-Severe was included in the network via its historical control^31^, results were similar to the base case analysis (**Figure S12**).

## Discussion

Clinical studies^8^, along with recent real-world evidence^8,33,34^, have demonstrated a mortality benefit for remdesivir in patients hospitalized with COVID-19 requiring supplemental oxygen. Our network meta-analysis demonstrates that among patients receiving low-flow oxygen, treatment with remdesivir consistently improved clinical outcomes including lowering the risk of mortality, improving recovery, increasing the likelihood of no longer requiring oxygen support and lowering the risk of progression to NIV or worse; results were similar when excluding the ACTT-2 trial. In patients treated with remdesivir in combination with baricitinib, the magnitude of effect was higher, indicating potentially synergistic effects, particularly in the high-flow group. These results support the conditional approval by the EMA and multiple jurisdictions globally that have recommended remdesivir for the treatment of COVID-19 patients.^19,35,36^

As observed in clinical practice^37^, and supported by the results of this meta-analysis, the effect of remdesivir on clinical outcomes varies depending on the degree of respiratory support at baseline. This meta-analysis suggests that the degree of respiratory support may be a useful indicator for treatment decisions. However, optimized surrogate markers for disease progression, including the identification of the pathophysiologic stages of COVID-19^38,39^ and the biological plausibility of the association between viral replication and pathophysiologic processes, are needed to further understand the clinical benefit of phase-specific treatments in COVID-19.

We also found that remdesivir in combination with baricitinib was superior to remdesivir monotherapy across all endpoints: the combination of an antiviral (remdesivir) with an anti-inflammatory (such as baricitinib, corticosteroids, or tocilizumab), as recommended in the National Institute of Health guidelines for the treatment of COVID-19, may be an effective treatment strategy for COVID-19 and should be further assessed.^40^ While treatment with remdesivir monotherapy resulted in significant improvements in mortality, recovery and progression among patients on low flow oxygen, the presence of baricitinib increased the magnitude of benefit observed across all endpoints.

The results of this meta-analysis differ from previous studies due to various reasons. Prior meta-analyses that have assessed the efficacy of remdesivir have included studies evaluating patients with heterogenous severity of COVID-19 disease or when less RCT evidence was available.^10-13,15-20^ In situations where meta-analyses were used to inform guideline recommendations^41^, imprecision in severity assessment may have compromised the validity of the recommendation.^42^ Other key differences include that prior meta-analyses included a more variable, smaller, study sample, in some cases without regard to receipt of supplemental oxygen at baseline. For example, remdesivir’s impact on mortality reported by the WHO in the SOLIDARITY publication did not reach statistical significance in the overall population.^10^ However, in the subgroup of patients with low- and high-flow supplemental oxygen/non-mechanical ventilation, there was a numerical trend towards benefit of treatment with remdesivir, with 28-day mortality lower among those treated with remdesivir (9.4%) versus standard of care (10.6%).^10^ Our results, when exploring low- and high-flow supplemental oxygen separately, have shown a more pronounced benefit for remdesivir in the low-flow oxygen population as observed elsewhere.^21,42^ The lack of observed clinical benefit in the high-flow oxygen population may indicate that clinical benefit of remdesivir is most pronounced in patients receiving low-flow oxygen; however, observed differences may also be due to smaller sample size in the high-flow oxygen population and the inclusion of patients on NIV in the high-flow oxygen population in some studies, which may have confounded the results. Prior analyses generally considered treatment with remdesivir separately as 5-day or 10-day courses, versus aggregate treatment as in our analysis. As noted in the methods, prior analyses identified no difference in 5-versus 10-day treatment^13,25,26^; further, treatment up to 10 days has been recommended in clinical practice.^43^ Differences in heterogeneity of the standard of care arm and in reporting may prevent meaningful comparisons in certain cases; the impact of these differences on results is difficult to ascertain. Further methodological differences may also explain the differences observed in results. Previous analyses have differentially reported outcomes as odds ratios^15,17,18,41,44^, versus risk ratios in our analysis, which only approximate each other when event rates are low, which is not the case for all endpoints. The Cochrane review did not consider the proportion of patients who recovered, but looked at time to recovery and determined these data were not able to be synthesized; therefore, recovery was not assessed in their meta-analysis.^20^ Other analyses, such as the meta-analysis for mortality published alongside the SOLIDARITY trial, have drawn conclusions regarding statistical significance based on 99% confidence intervals,^42^ as opposed to the more standard 95% intervals employed in our analysis. Further, given their large sample size and contribution to the network, this confounding factor may bias the results of any meta-analysis that includes this data.

To the best of our knowledge, this is the first meta-analysis performed in remdesivir’s EMA-indicated population that incorporates patient-level data from SIMPLE-Moderate. The similar model fits and results across the fixed and random effect models underlines the consistency and robustness of our results. However, the evaluation and synthesis of evidence in a rapidly evolving field is inherently associated with limitations. First, SIMPLE-Severe could not connect to a network in the base case analysis as it compared 5-versus 10-day treatments of remdesivir (with no further control arm)^25^; however, a scenario analysis where it was included through its historical control did not meaningfully impact the results. Second, the heterogeneity of the included trials may limit the generalizability of the results. For example, SOLIDARITY did not require all patients to have a confirmed infection of COVID-19, and the inclusion of patients was left at the discretion of the enrolling physician; further, the protocol exclusion criteria were ambiguous. For reasons unknown, mortality rates observed in SOLIDARITY’s best supportive care arm were higher than those observed across other studies conducted in a similar time period. Given SOLIDARITY’s large sample size, these limitations may contribute disproportionately to the results of this analysis. Third, this meta-analysis excludes the recent results of the ACTT-3^45^ and the DisCoVeRy trial^46^, both of which were published after our search. While ACTT-3 showed similar effects to studies included in this meta-anlsysis of remdesivir alone on mortality rates, DisCoVeRy was a sub-study of SOLIDARITY and the DisCoVeRy trial would have been excluded to avoid potential bias due to double-counting patients. Fourth, across our included trials, the definition of recovery varied and for the purpose of synthesizing our evidence, we assumed discharge to be equivalent to recovery where recovery was not reported as a distinct outcome. Fifth, we assumed that outcomes reported at day 24 were equivalent to those reported at day 28 in the analysis. Sixth, the trials included enrolled patients from across multiple geographic regions with varying definitions of best supportive care that have evolved since the beginning of the pandemic; these differences have likely impacted mortality not only between regions but also over time, as evidence emerges on best supportive care for patients with COVID-19. Finally, the data informing our meta-analysis was identified through a targeted, rather than a systematic, literature review. However, given the constrained nature of the disease area and the ability to extensively validate the included studies using other recently conducted meta-analyses, this is likely not a limitation.

In patients with COVID-19 requiring any or low-flow supplemental oxygen at baseline, based on available randomized clinical trial evidence, this analysis found that treatment with remdesivir lowered mortality, accelerated recovery and reduced progression to NIV, compared to best supportive care.

Future studies exploring the impact of antivirals, notably baricitinib, in patients may provide additional data to explain these findings. The results of this study suggest that remdesivir should be considered as part of a multi-faceted care strategy for these patients.

## Data Availability

All data produced in the present work are contained in the manuscript.

## Author contributions

All authors contributed to the design of the research. RB, NS and SJ performed the analysis of the results with all authors contributing to the discussion and interpretation of the results. RB took the lead in writing the manuscript, with all authors providing critical feedback on all drafts of the manuscript.

