## Supplementary Materials for "Remdesivir for the treatment of patients hospitalized with COVID-19 receiving supplemental oxygen: a targeted literature review and meta-analysis"

**Table S1.** PRISMA checklist: the preferred reporting items for systematic reviews and meta-analyses^21^

|  | **Section/topic** | | **#** | **Checklist item** | | **Reported on page #** |
| --- | --- | --- | --- | --- | --- | --- |
|  | **Title** | | | | |  |
|  | Title | | 1 | Identify the report as a systematic review, meta-analysis, or both. | | 1 |
|  | **Abstract** | | | | |  |
|  | Structured summary | | 2 | Provide a structured summary including, as applicable: background; objectives; data sources; study eligibility criteria, participants, and interventions; study appraisal and synthesis methods; results; limitations; conclusions and implications of key findings; systematic review registration number. | | 2 |
|  | **Introduction** | | | | |  |
|  | Rationale | | 3 | Describe the rationale for the review in the context of what is already known. | | 3-4 |
|  | Objectives | | 4 | Provide an explicit statement of questions being addressed with reference to participants, interventions, comparisons, outcomes, and study design (PICOS). | | 3-4 |
|  | **Methods** | | | | |  |
|  | Protocol and registration | | 5 | Indicate if a review protocol exists, if and where it can be accessed (e.g., Web address), and, if available, provide registration information including registration number. | | N/A |
|  | Eligibility criteria | | 6 | Specify study characteristics (e.g., PICOS, length of follow-up) and report characteristics (e.g., years considered, language, publication status) used as criteria for eligibility, giving rationale. | | 4 |
|  | Information sources | | 7 | Describe all information sources (e.g., databases with dates of coverage, contact with study authors to identify additional studies) in the search and date last searched. | | 4  Supplementary Materials |
|  | Search | | 8 | Present full electronic search strategy for at least one database, including any limits used, such that it could be repeated. | | Supplementary Materials |
|  | Study selection | | 9 | State the process for selecting studies (i.e., screening, eligibility, included in systematic review, and, if applicable, included in the meta-analysis). | | 4 |
|  | Data collection process | | 10 | Describe method of data extraction from reports (e.g., piloted forms, independently, in duplicate) and any processes for obtaining and confirming data from investigators. | | 4 |
|  | Data items | | 11 | List and define all variables for which data were sought (e.g., PICOS, funding sources) and any assumptions and simplifications made. | | 4-5 |
|  | Risk of bias in individual studies | | 12 | Describe methods used for assessing risk of bias of individual studies (including specification of whether this was done at the study or outcome level), and how this information is to be used in any data synthesis. | | 4-5 |
|  | Summary measures | | 13 | State the principal summary measures (e.g., risk ratio, difference in means). | | 5 |
|  | Synthesis of results | | 14 | Describe the methods of handling data and combining results of studies, if done, including measures of consistency (e.g., I^2^) for each meta-analysis. | | 5 |
|  | Risk of bias across studies | | 15 | Specify any assessment of risk of bias that may affect the cumulative evidence (e.g., publication bias, selective reporting within studies). | | N/A |
|  | Additional analyses | | 16 | Describe methods of additional analyses (e.g., sensitivity or subgroup analyses, meta-regression), if done, indicating which were pre-specified. | | 5 |
| **Results** | | | | | |  |
| Study selection | | 17 | | | Give numbers of studies screened, assessed for eligibility, and included in the review, with reasons for exclusions at each stage, ideally with a flow diagram. | 6 |
| Study characteristics | | 18 | | | For each study, present characteristics for which data were extracted (e.g., study size, PICOS, follow-up period) and provide the citations. | Table 1 - 2 |
| Risk of bias within studies | | 19 | | | Present data on risk of bias of each study and, if available, any outcome level assessment (see item 12). | Table S5 |
| Results of individual studies | | 20 | | | For all outcomes considered (benefits or harms), present, for each study: (a) simple summary data for each intervention group (b) effect estimates and confidence intervals, ideally with a forest plot. | Table 3 |
| Synthesis of results | | 21 | | | Present the main results of the review. If meta-analyses are done, include for each, confidence intervals and measures of consistency. | 7 |
| Risk of bias across studies | | 22 | | | Present results of any assessment of risk of bias across studies (see Item 15). | N/A |
| Additional analysis | | 23 | | | Give results of additional analyses, if done (e.g., sensitivity or subgroup analyses, meta-regression [see Item 16]). | 8 |
| **Discussion** | | | | | |  |
| Summary of evidence | | 24 | | | Summarize the main findings including the strength of evidence for each main outcome; consider their relevance to key groups (e.g., healthcare providers, users, and policy makers). | 9 |
| Limitations | | 25 | | | Discuss limitations at study and outcome level (e.g., risk of bias), and at review-level (e.g., incomplete retrieval of identified research, reporting bias). | 10 - 11 |
| Conclusions | | 26 | | | Provide a general interpretation of the results in the context of other evidence, and implications for future research. | 10 - 11 |
| **Funding** | | | | | |  |
| Funding | | 27 | | | Describe sources of funding for the systematic review and other support (e.g., supply of data); role of funders for the systematic review. | 1 |

**Table S2.** Search strategy^a^

| **Search Terms** | | |
| --- | --- | --- |
| **Intervention terms** | **1** | Remdesivir [All fields] |
| **Study type** | **2** | “Randomized controlled trial” or RCT or “clinical study” or “clinical trial” or randomization [All fields] |
|  | **3** | “clinical study” or “clinical trial” or “clinical trial, Phase I” or “clinical trial, Phase II” or “clinical trial, Phase III” or “Clinical trial, Phase IV”, or “comparative study” or “controlled clinical trial" or "pragmatic clinical trial" or "randomized controlled trial" [Publication Type] |
|  | **4** | 1 AND (2 OR 3) |
|  | **5** | 1 AND 2 |

.

**Table S3.** Priors implemented by default in BUGSnet (adapted from Béliveau et al.^28^)

| Parameters | Consistency Model | |
| --- | --- | --- |
|  | Random effect | Fixed effect |
| **µ_1, …,_ µ*_M_*** | iid N(0,(15*u*)^2^) |  |
|  | Except when a log link is used with a binomial family, in which case:  *µ_I_* = log(*p_i_*), *p_i_* ~iid U(0,1) | |
| ***d*_1, 2, …,_ *d*_1,_ *_T_*** | iid N(0,(15*u*)^2^) |  |
| ***d*_1, 2,_ …, *d*_1,_ *_T,_ …,***  ***d_T_* _– 2,_ *_T_* _– 1_, *d_T_* _– 2,_ *_T_*, *d_T_* _– 1,_ *_T_*** | NA |  |
| ***σ*** | U(0,*u*) | NA |
| **β_(1,2)_, …, β_(1,_ *_K_*_)_**  **(meta-regression only)** | Unrelated: iid t(0, *u*^2^, df = 1)  Exchangeable: iid N(*b*, γ^2^), *b*~t(0, *u*^2^, df=1), γ~*U*(0, *u*)  Equal: β_2_= … = β*_T_ = B, B*~ t(0, *u*^2^, df=1) | |

**Table S4.** Model fit - fixed effects versus random effects

| Endpoint | Time | Subgroup | Fixed Effects Model | | | Random Effects Model | | |
| --- | --- | --- | --- | --- | --- | --- | --- | --- |
|  |  |  | pD | Dres | DIC | pD | Dres | DIC |
| **Mortality** | Early | *Any Flow* | NC | 13.86 | NC | NC | 9.05 | NC |
|  |  | *High Flow* | NC | 5.85 | NC | NC | 5.81 | NC |
|  |  | *Low Flow* | NC | 7.77 | NC | NC | 6.24 | NC |
|  | Later | *Any Flow* | NC | 19.44 | NC | NC | 14.86 | NC |
|  |  | *High Flow* | NC | 5.7 | NC | NC | 5.68 | NC |
|  |  | *Low Flow* | NC | 7.91 | NC | NC | 6.21 | NC |
| **Recovery** | Early | *Any Flow* | 6.97 | 20.87 | 27.83 | 8.97 | 9.7 | 18.67 |
|  |  | *High Flow* | NC | 7.39 | NC | NC | 6.2 | NC |
|  |  | *Low Flow* | 4.89 | 7.02 | 11.91 | 5.57 | 6.11 | 11.68 |
|  | Later | *Any Flow* | 7.78 | 15.63 | 23.42 | 9.78 | 11.22 | 21 |
|  |  | *High Flow* | NC | 7.39 | NC | NC | 6.5 | NC |
|  |  | *Low Flow* | 4.68 | 6.03 | 10.71 | 5.23 | 5.9 | 11.14 |
| **Free from oxygen support** | Early | *Any Flow* | 5.86 | 6.81 | 12.67 | 6.63 | 7.18 | 13.81 |
|  |  | *High Flow* | NC | 7.63 | NC | NC | 6.27 | NC |
|  |  | *Low Flow* | 4.71 | 7.05 | 11.76 | 5.42 | 6.17 | 11.58 |
| **Requiring NIV or worse** | Early | *Any Flow* | 6.9 | 13.76 | 20.65 | 9.15 | 11.04 | 20.19 |
|  |  | *High Flow* | 4.64 | 4.99 | 9.62 | 4.83 | 5.16 | 9.99 |
|  |  | *Low Flow* | NC | 12.91 | NC | NC | 6.06 | NC |
| **Requiring IMV or worse** | Early | *Any Flow* | NC | 15.24 | NC | NC | 8.77 | NC |
|  |  | *High Flow* | NC | 5.65 | NC | NC | 5.63 | NC |
|  |  | *Low Flow* | NC | 11.51 | NC | NC | 6.11 | NC |

DIC: deviance information criterion; Dres: posterior mean of the residual deviance; IMV: invasive mechanical ventilation; NC: not computable; NIV: non-invasive ventilation; pD: effective number of parameters

**Table S5.** Risk of bias assessment ^23^

| Study | Randomization process | | Bias due to deviations from intended interventions | | Bias due to missing outcome^a^ data | | Bias in measurement of the outcome^a^ | | Bias in selection of reported results | | Overall bias |
| --- | --- | --- | --- | --- | --- | --- | --- | --- | --- | --- | --- |
| ACTT-1^8^ |  | |  | |  | |  | |  | |  |
| ACTT-2^9^ |  | |  | |  | |  | |  | |  |
| Hubei^31^ |  | |  | |  | |  | |  | |  |
| SOLIDARITY^10^ |  | |  | |  | |  | |  | |  |
| SIMPLE-Moderate^25^ |  | |  | |  | |  | |  | |  |
| Mahajan^22^ |  | |  | |  | |  | |  | |  |
|  | Low risk of bias |  | | High risk of bias | |  | | Some concerns | |  |  |

^a^Assessed for primary outcome

**Table S6.** Fixed effects SUCRA findings

| Endpoint | Time | Subgroup | RDV | BAR+RDV | BSC |
| --- | --- | --- | --- | --- | --- |
| **Mortality** | Early | *Any Flow* | 12.7% | 87.3% | 0.0% |
|  |  | *High Flow* | 10.8% | 86.5% | 2.7% |
|  |  | *Low Flow* | 47.5% | 52.5% | 0.0% |
|  | Later | *Any Flow* | 2.0% | 98.0% | 0.0% |
|  |  | *High Flow* | 10.4% | 78.8% | 10.7% |
|  |  | *Low Flow* | 5.7% | 94.4% | 0.0% |
| **Recovery** | Early | *Any Flow* | 0.3% | 99.4% | 0.3% |
|  |  | *High Flow* | 0.1% | 99.2% | 0.7% |
|  |  | *Low Flow* | 15.7% | 84.3% | 0.0% |
|  | Later | *Any Flow* | 0.4% | 99.6% | 0.0% |
|  |  | *High Flow* | 0.2% | 82.4% | 17.5% |
|  |  | *Low Flow* | 10.3% | 89.7% | 0.0% |
| **Free from oxygen support** | Early | *Any Flow* | 0.0% | 100.0% | 0.0% |
|  |  | *High Flow* | 0.0% | 100.0% | 0.0% |
|  |  | *Low Flow* | 3.4% | 96.6% | 0.0% |
| **Requiring NIV or worse** | Early | *Any Flow* | 0.2% | 99.9% | 0.0% |
|  |  | *High Flow* | 0.1% | 99.8% | 0.1% |
|  |  | *Low Flow* | 26.7% | 73.4% | 0.0% |
| **Requiring IMV or worse** | Early | *Any Flow* | 0.0% | 100.0% | 0.0% |
|  |  | *High Flow* | 1.0% | 98.6% | 0.4% |
|  |  | *Low Flow* | 1.8% | 98.2% | 0.0% |

BAR: baricitinib; BSC: best supportive care; IMV: invasive mechanical ventilation; NIV: non-invasive ventilation; RDV: remdesivir

**Figure S1.** Network diagram plots, base case analysis, by time point assessment and type of non-invasive oxygen support at baseline. (A) Early, any flow oxygen, (B) Early, low-flow oxygen, (C) Early, high-flow oxygen, (D) Later, any flow oxygen, (E) Later, low-flow oxygen and (F) Later, high-flow oxygen.

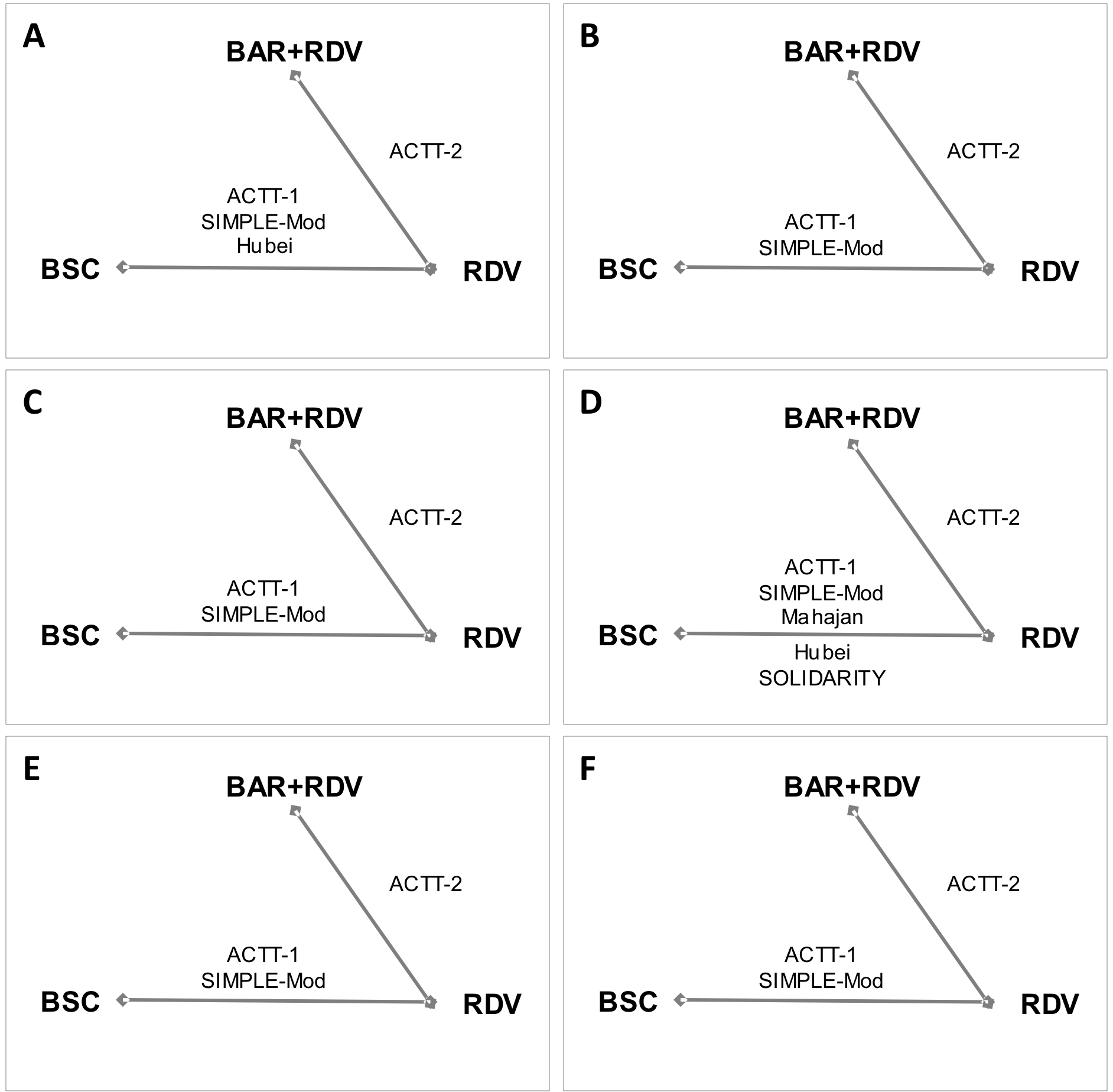

BAR: baricitinib; BSC: best supportive care; IMV: invasive mechanical ventilation; NIV: non-invasive ventilation; RDV: remdesivir

**Figure S2**. Posterior mean deviance comparison for consistency versus inconsistency model for mortality by time point assessment and type of non-invasive oxygen support at baseline. (A) Early, any flow oxygen, (B) Early, low-flow oxygen, (C) Early, high-flow oxygen, (D) Later, any flow oxygen, (E) Later, low-flow oxygen and (F) Later, high-flow oxygen.
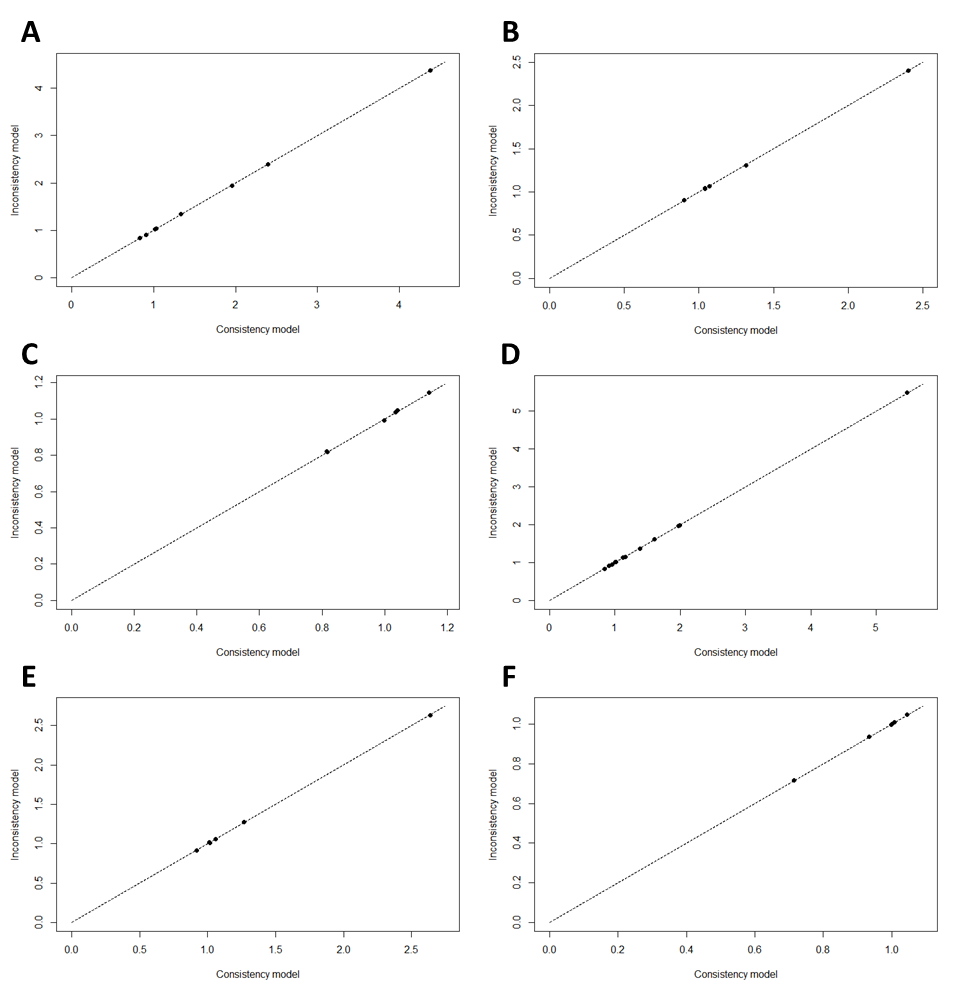

**Figure S3**. Posterior mean deviance comparison for consistency versus inconsistency model for recovery by time point assessment and type of non-invasive oxygen support at baseline. (A) Early, any flow oxygen, (B) Early, low-flow oxygen, (C) Early, high-flow oxygen, (D) Later, any flow oxygen, (E) Later, low-flow oxygen and (F) Later, high-flow oxygen.
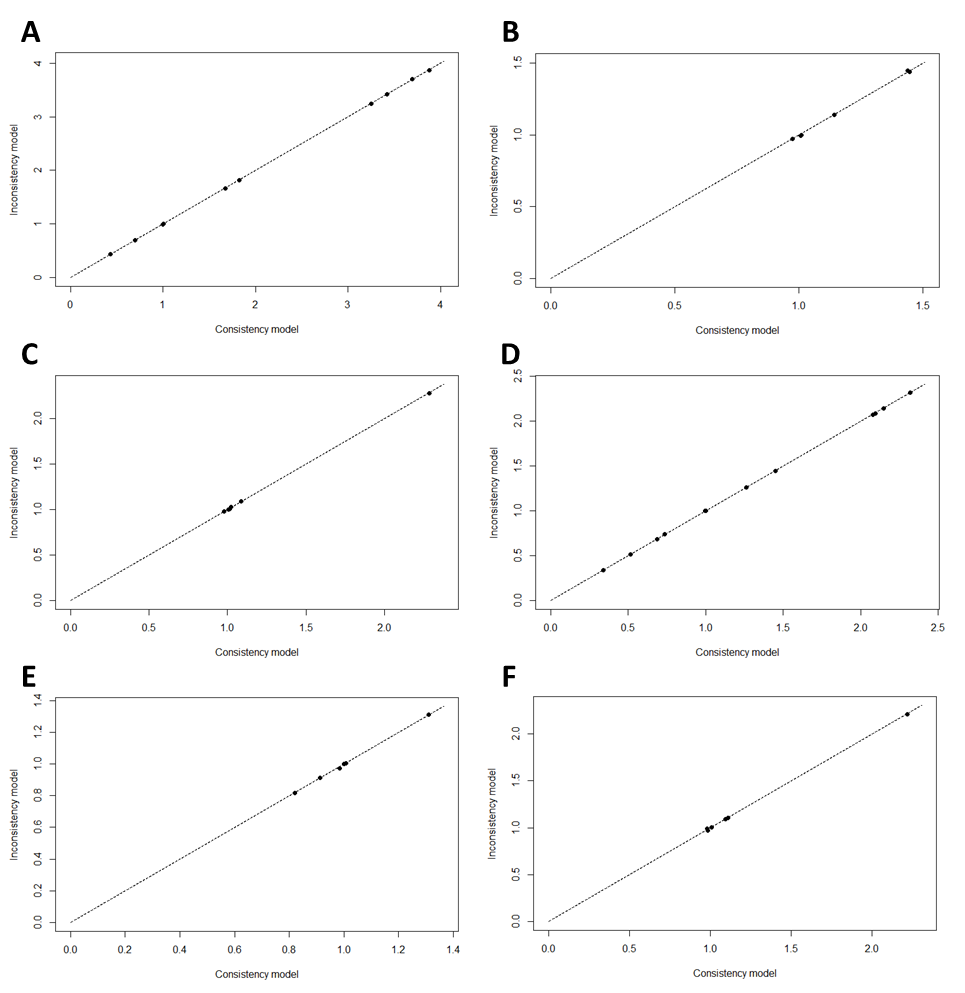

**Figure S4**. Posterior mean deviance comparison for consistency versus inconsistency model for no longer requiring oxygen support by type of non-invasive oxygen support at baseline. (A) Any flow oxygen, (B) Low-flow oxygen, (C) High-flow oxygen.
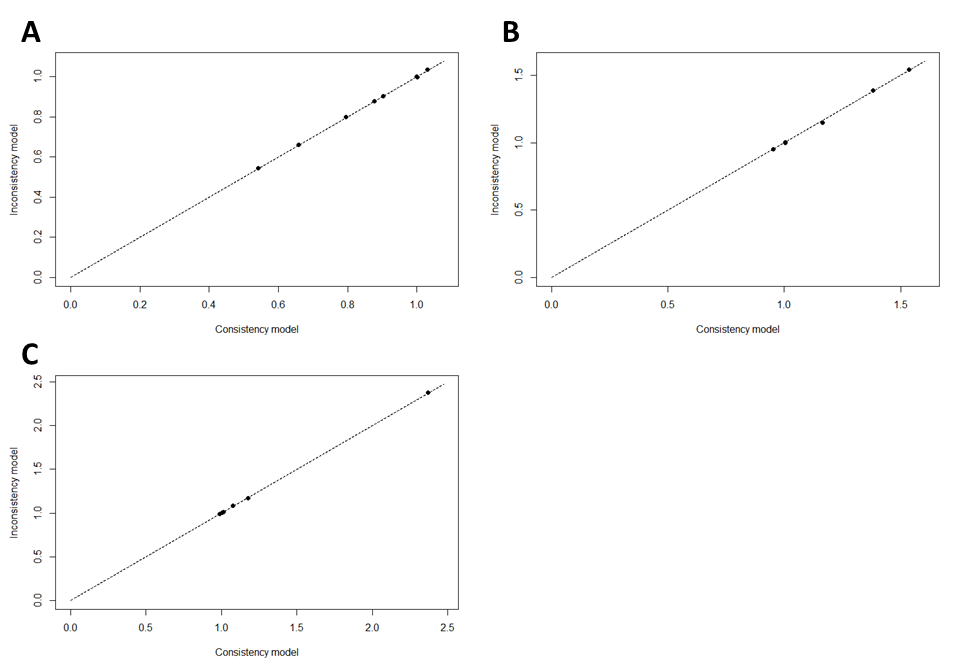

**Figure S5**. Posterior mean deviance comparison for consistency versus inconsistency model for requiring non-invasive ventilation or worse by type of non-invasive oxygen support at baseline. (A) Any flow oxygen, (B) Low-flow oxygen, and (C) High-flow oxygen.
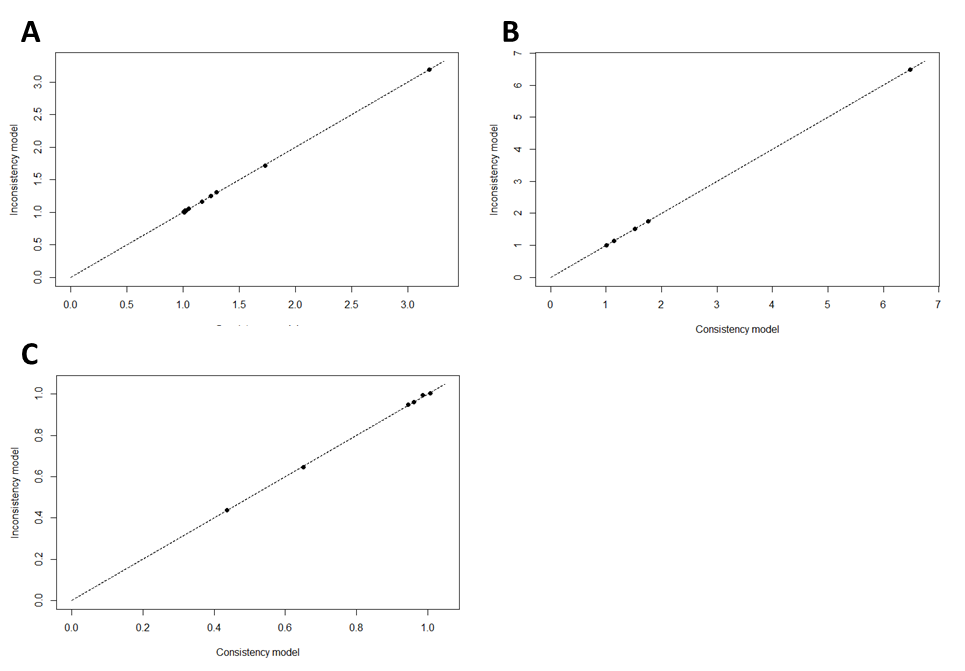

**Figure S6**. Posterior mean deviance comparison for consistency versus inconsistency model for requiring invasive ventilation or worse by type of non-invasive oxygen support at baseline. (A) Any flow oxygen, (B) Low-flow oxygen, and (C) High-flow oxygen
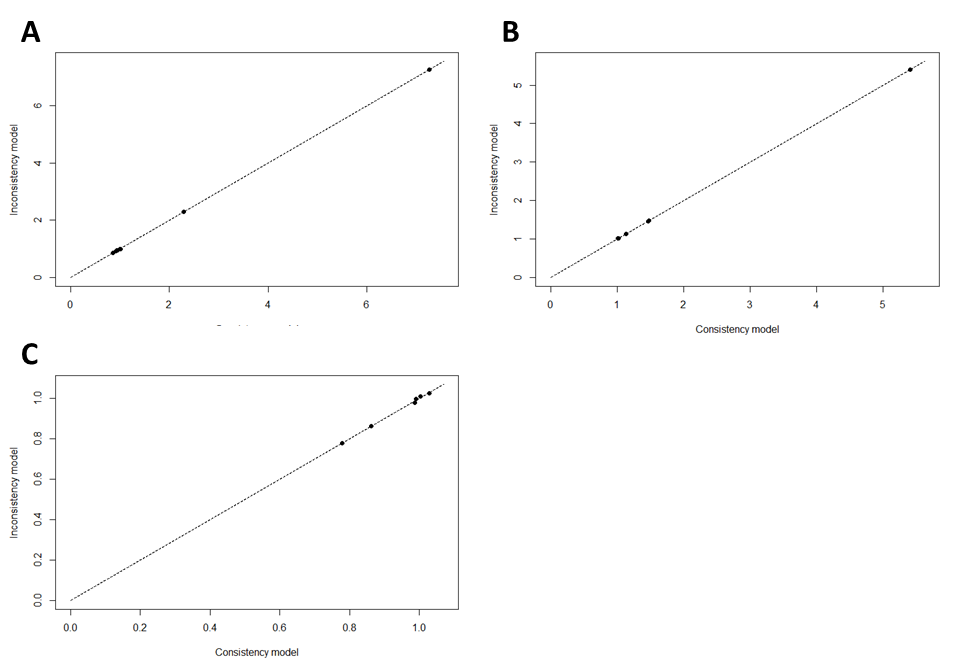

**Figure S7.** Scenario analysis of disaggregated 5- and 10-day remdesivir treatment: forest plot for mortality endpoint at early time assessment only, by type of non-invasive oxygen support at baseline

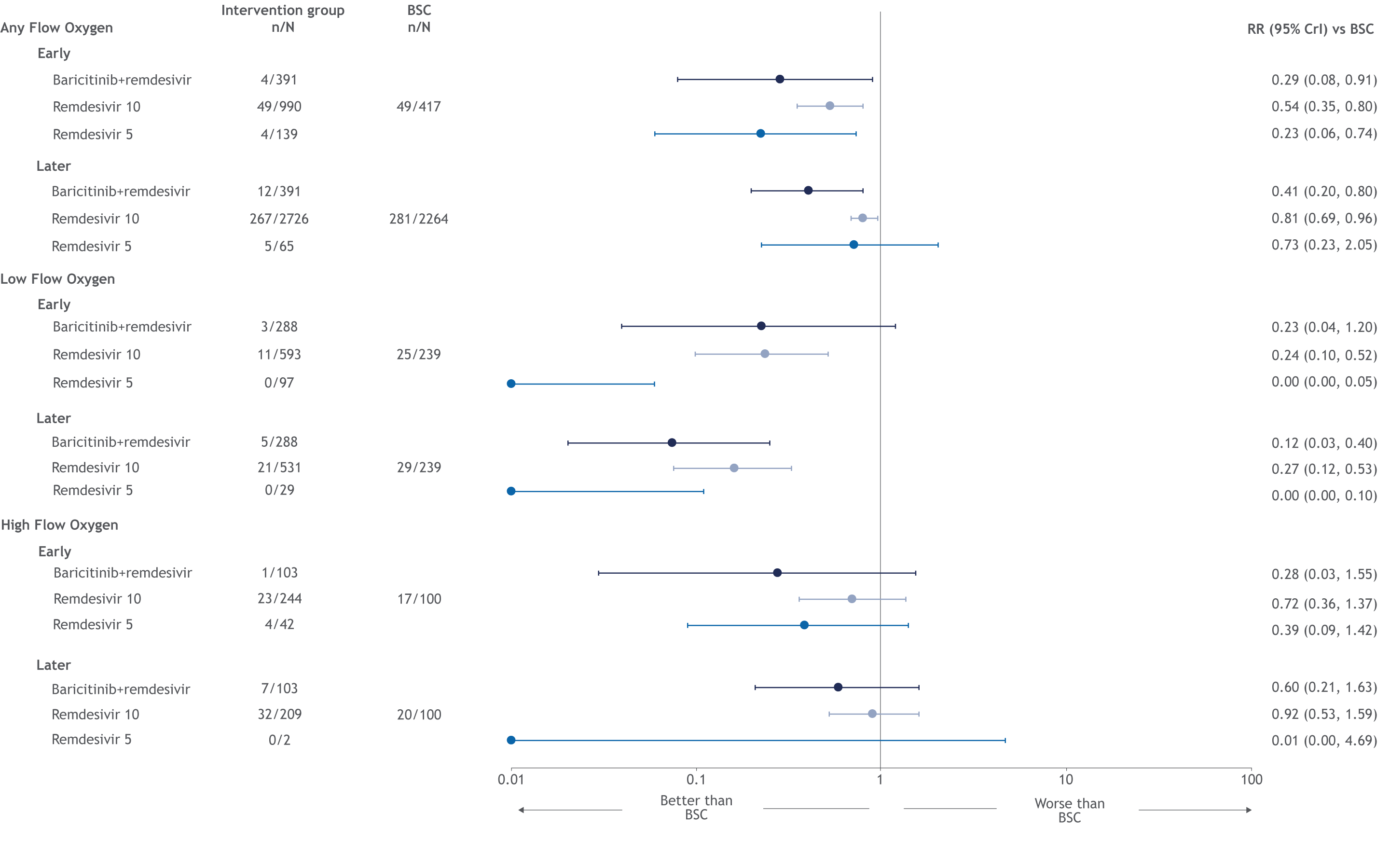
BSC: best supportive care; RR: risk ratios

**Figure S8.** Scenario analysis of excluding treatment with baricitinib: forest plot for mortality endpoint at early time assessment only, by type of non-invasive oxygen support at baseline

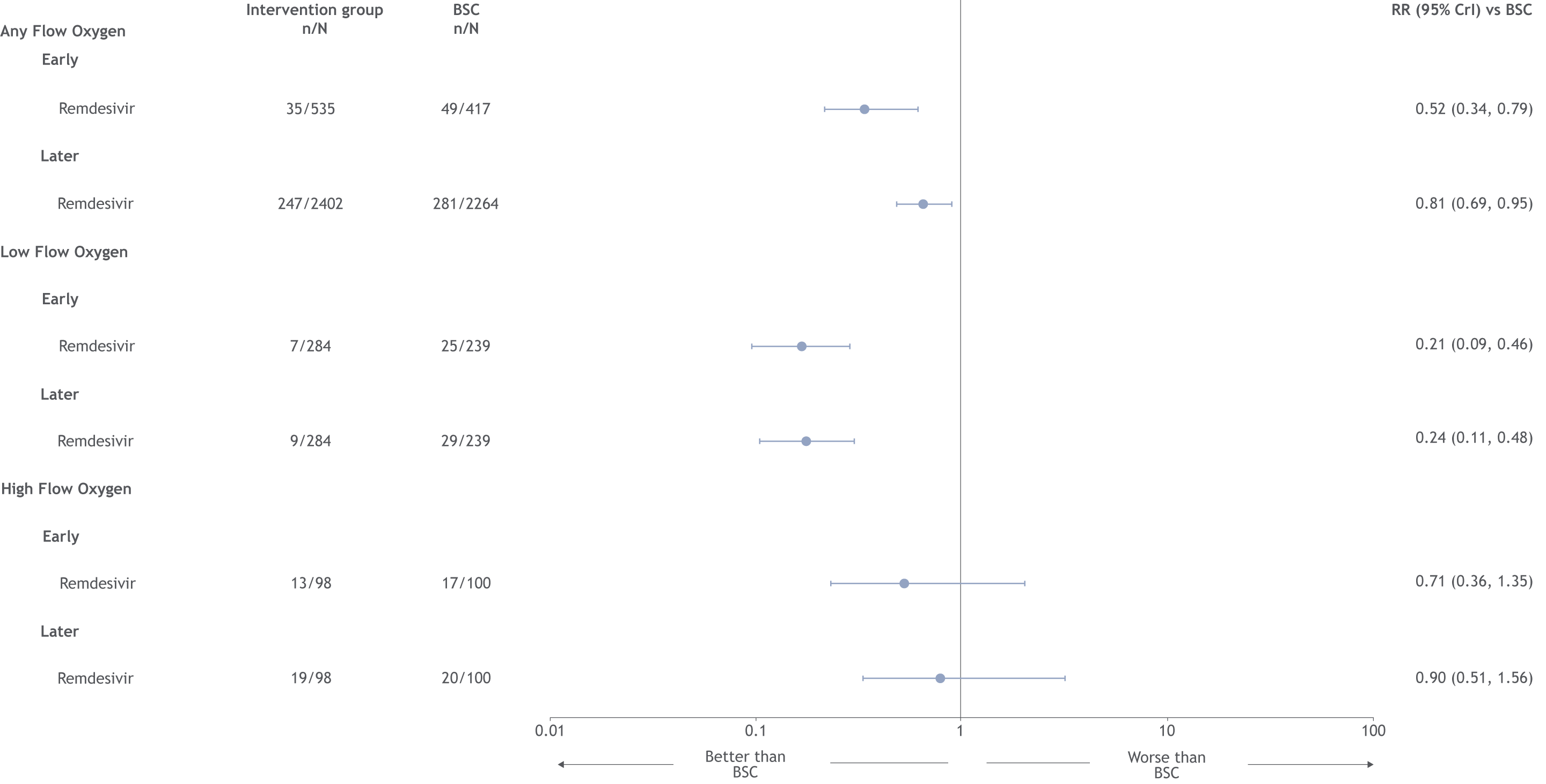

BSC: best supportive care; RR: risk ratios

**Figure S9.** Scenario analysis of excluding treatment with baricitinib: forest plot for recovery endpoint at early time assessment only, by type of non-invasive oxygen support at baseline

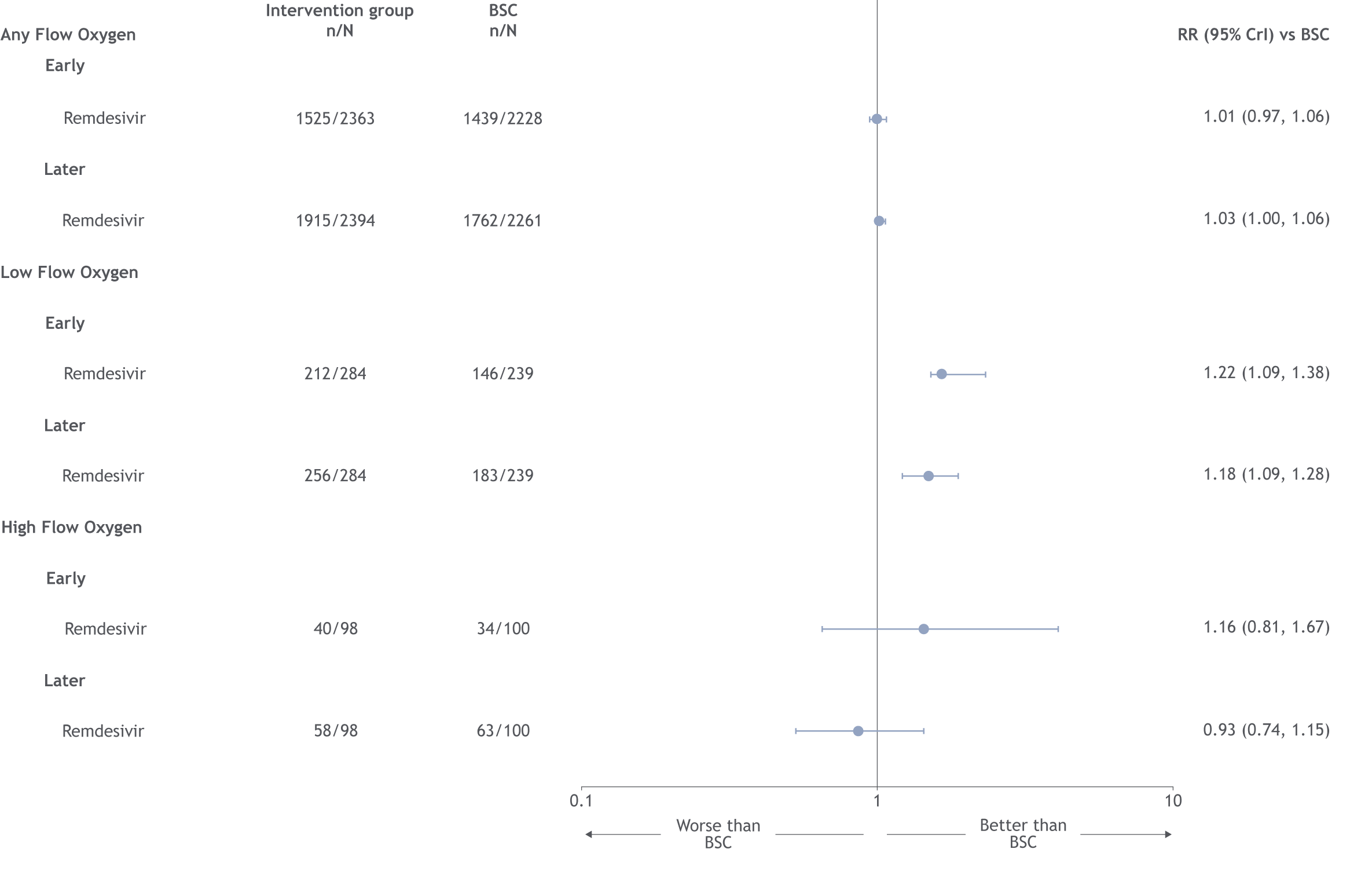

BSC: best supportive care; RR: risk ratio

**Figure S10.** Scenario analysis of excluding treatment with baricitinib: forest plot for no longer requiring oxygen endpoint at early time assessment only, by type of non-invasive oxygen support at baseline

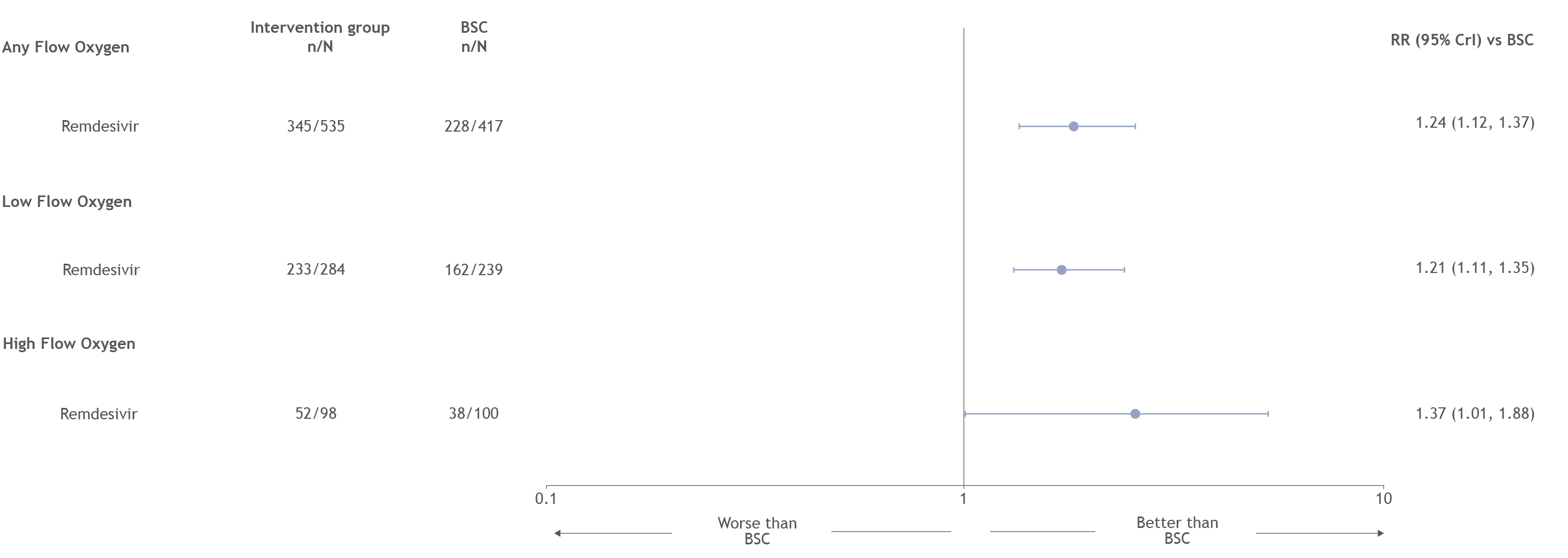

BSC: best supportive care; RR: risk ratio

**Figure S11.** Scenario analysis of excluding treatment with baricitinib: forest plot for NIV or IMV or worse endpoint at early time assessment only, by type of non-invasive oxygen support at baseline

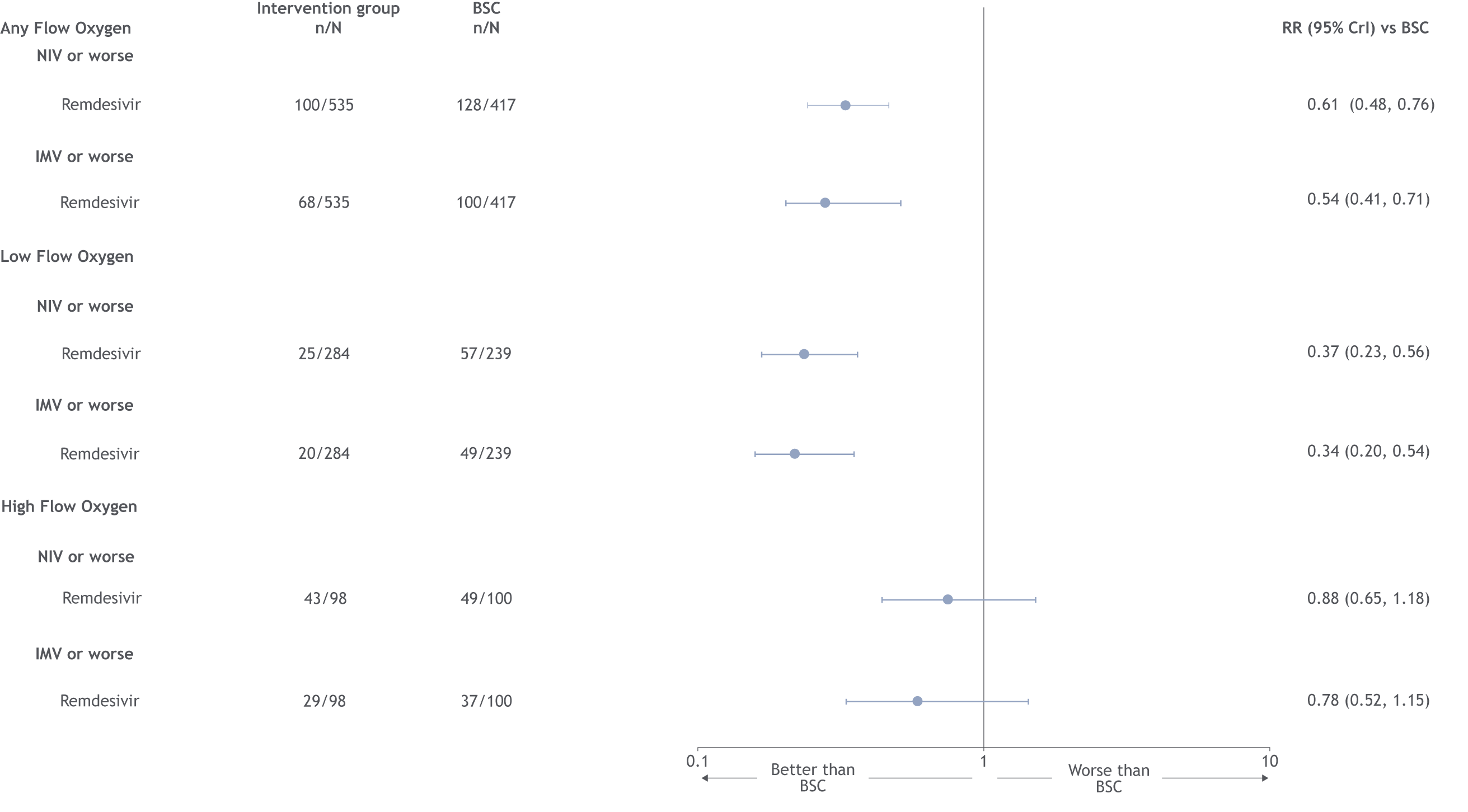

BSC: best supportive care; IMV: invasive mechanical ventilation; NIV: non-invasive ventilation; RR: risk ratio
**Figure S12.** Scenario analysis of inclusion of SIMPLE-Severe^24^: forest plot for mortality endpoint at early time assessment only, by type of non-invasive oxygen support

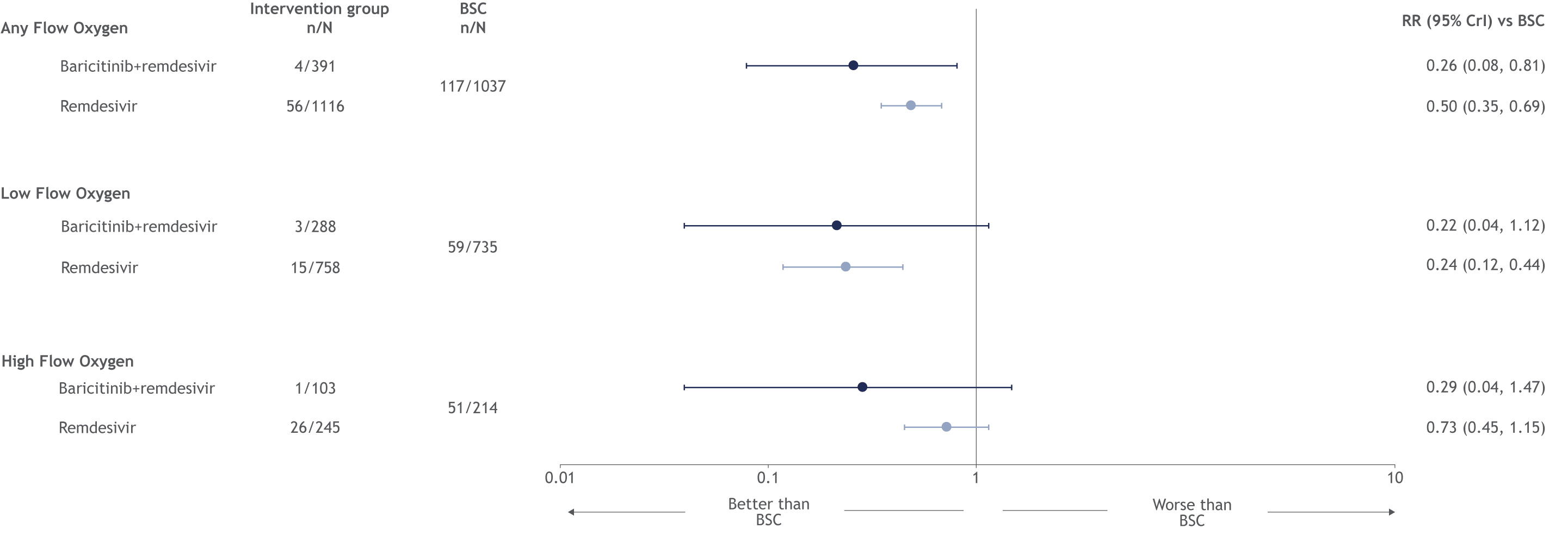

BAR: baricitinib; BSC: best supportive care; RDV: remdesivir; RR: risk ratios
